# Crowdsourcing AI solutions in early rehospitalisation as a pilot for healthcare regulatory sandbox

**DOI:** 10.64898/2026.09.10.26362629

**Authors:** Miha Keber, Moris Bagić, Ilona Kulikovskikh, Matija Piškorec, Pero Ivanko, Davor Oršolić, Milica Projić, Željana Barišić, Ana Jerončić, Nina Šesto, Maša Šams-Bival, Siniša Košćina, Zrinka Potočanac, Andrija Štajduhar, Jan Kolić, Karlo Pintarić, Lovro Trgovec-Greif, Martin Trgovec-Greif, Iva Buljan, Roko Šango, Filip Miočinović, Filip Ðerke, Zoran Antolović, Milan Pavlović, Tomislav Šmuc, Anja Barešić

## Abstract

The uptake of new digital and AI based technologies in the healthcare setting, including digital diagnostics, has been comparatively slower than in other sectors. This is a consequence of the complex legislature, and the lack of routine procedures for the secondary use of sensitive medical data. Inadequate level of education and incentives for different stakeholders prevents overcoming strong barriers, such as clinical workflow integration, and need for comprehensive evaluation of utility of interventions which requires multi-disciplinary collaboration effort. The paper presents a re-hospitalization challenge, organised by the European digital innovation hub (EDIH) - AI4Health.Cro, a co-creation activity and an effective tool for overcoming the mentioned barriers for implementation of new AI technologies and overall innovation ecosystem maturation. Crowdsourcing challenges or datathons are R&D strategies where ad-hoc assembled competing teams collaborate on early-stage technology development or research problem solving. Our re-hospitalisation challenge has had multiple facets: as activity for construction of an AI predictive model with explanatory support and plausible business model for its deployment; it involved also extensive organisers’ effort: data anonymisation, provision of secure processing environment, and comprehensive assessment of solutions. The paper details the challenge structure and processes, from problem inception to execution and evaluation. As a co-creation effort in digital healthcare, the challenge provided results on multiple levels: (i) tailored solutions based on primary data, serving as prototypes for later deployment in practice; (ii) sandboxing EHDS principles to enable secondary data use; (iii) ecosystem building event yielding cross-disciplinary participative learning experience. Innovation challenges aren’t just competitions; they are a building tool that provides several direct and indirect impacts, from developing healthcare technologies, supporting startups, to streamlining secondary use of clinical data for research and innovation and eliciting regulatory needs. Raising the awareness of all relevant stakeholders about the translation of AI technologies into routine clinical setting.

**Author summary:** Healthcare has been slower than other sectors to adopt new digital and AI-based technologies. This slow progress largely reflects the complexity of EU legislation and the lack of practical procedures that translate legal requirements into guidance for stakeholders using sensitive medical data in research and innovation. A crowdsourcing pilot to predict likelihood of early rehospitalisation using AI methodology provided opportunity for hands-on experience for experts from clinical, computational and business background on both synthetic and real-world medical data. Together we clarify legal responsibilities and data access conditions, and facilitate collaboration of relevant stakeholders, clinical, technical, public and research entities for ecosystem building. We provide the first Croatian European Health Data Space compliant secure processing environment, created for future use by researchers and innovators. Methodologically varied AI based prototypes were developed to predict early rehospitalization carefully balancing data overfitting, AI explainability and market potential of technology, in turn raising multidisciplinary skills, and awareness of conditions required for innovation to thrive in 28 participating teams. This type of competition, provided within European Digital Innovation Hub is an ideal platform to build regulatory sandboxes and pilot novel legislative platforms ahead of their full-scale implementation.

## Introduction

The increasing use of artificial intelligence (AI) in healthcare relies on access to sensitive private data, which must be handled under strict regulations such as General Data Protection Regulation (GDPR) and the European Health Data Space (EHDS) to ensure patient privacy and trust [1]. Integrating AI-driven insights into the evidence-based medicine framework ensures that clinical expertise and patient needs remain central to responsible medical decision-making. Crowdsourcing computational challenges, such as healthcare datathons, harness diverse expertise to develop innovative AI solutions while maintaining compliance with these frameworks. In this work we describe organisation and execution of a crowdsourcing competition, we refer to it as the challenge, around the problem of predicting re-hospitalisation. The technical aim of the challenge was to develop effective digital tool that would help clinicians [2] in identification of high-risk patients with respect to 30-day re-hospitalisation time period, in order to help improve continuity of care, and reduce unnecessary readmissions. A number of other objectives associated to the challenge as a strategic tool driving digital transformation and healthcare innovation community [3, 4] and ecosystem development are further explained and discussed.

### Re-hospitalisation as a prediction problem

Early re-hospitalisation is a significant clinical challenge caused by post-discharge complications, poorly managed chronic conditions (e.g. diabetes or heart failure) and poor medication adherence. These and other interacting factors necessitate machine learning approaches capable of analysing complex clinical variables across large datasets.

Re-hospitalisation prediction typically focuses on post-discharge period of 30 days, using diagnostic, clinical, and occasionally socioeconomic data [5, 6]. However, readmission definitions lack field-wide consensus, with reported rates from electronic health records varying between 5.9 − 54.4% for specific patient populations and 7.7 − 23% for general populations [7]. Most re-hospitalisation predictions in healthcare are based on often used LACE variables: length-of-stay, acuity of admission, Charlson Co-morbidity Index (CCI) [8] score, emergency room visits in previous 6 months [9], however, we decided to investigate a broader context by including a number of other variables listed in (S1 Appendix).

Re-hospitalisation prediction problem is well suited for machine learning, due to a high number of records, with a wealth of descriptors and well defined label (target variable). In recent works re-hospitalisation problem was modelled by logistic regression, decision trees, random forest, and gradient boosting and neural networks (NN) [5, 10, 11], with the latter NN showing the highest evaluation scores but lacking explainability.

In the healthcare domain, model explainability plays a critical role in enabling trustworthy deployment of AI systems. In the context of re-hospitalisation prediction problem, most commonly used methods for explaining AI models were feature importances based on ensembles of decision trees and SHAP [12]. Feature importance analyses consistently highlight the CCI and similarly Elixhauser Co-morbidity Index [13] as most relevant features [5, 6, 10, 11]. In another study logistic regression [5] identified some of the most important variables to be hospitalisation through the emergency room, CCI, and ICD−10 category of disease.

### Regulatory frameworks and digital innovation ecosystem

To govern the deployment of the machine learning models discussed in the previous section, the European Union’s AI Act (AIA), effective as of July 2024, establishes essential safety and ethical standards. Under the AIA, predictive models for hospital readmission are classified as high-risk, requiring rigorous validation to protect patient outcomes while fostering innovation [14]. Complementing this, the EHDS facilitates the secure, standardised sharing of health data across member states. By utilising secure processing environments (SPEs), the EHDS allows developers to train robust, generalizable AI models on diverse datasets while maintaining strict compliance with privacy mandates [1]. As these AI solutions mature, they further intersect with the EU Medical Device Regulation for market approval and the Health Technology Assessment Regulation for reimbursement, both of which are critical for the large-scale deployment of digital medical devices [15].

While these regulations define the legal boundaries, European Digital Innovation Hubs (EDIHs) provide the practical infrastructure for digital transformation [16]. Functioning as “one-stop shops”, EDIHs offer technical consulting, pre-investment testing and and guidance on financing and regulatory compliance. In healthcare, EDIHs enable healthcare providers to safely experiment with AI-driven solutions and upskill staff, while reducing the risks associated with adopting complex digital tools. Within the EHDS framework, EDIHs play a vital complementary role as they provide the methodological support and expertise needed to validate and deploy AI solutions effectively.

### Study outline

In this work we detail the organisation, structure and management of an AI challenge on re-hospitalisation prediction problem. The challenge involved process of acquisition and preprocessing of data to make it appropriate for the purpose of the challenge. The challenge tasks and evaluation procedures moved beyond mere predictive performance, like explainability, usability, business aspects, bearing importance for the successful implementation of AI tools in the clinical setting. We further describe the implementation of a secure processing environment and the development of a credible anonymised dataset and its synthetic replica. In addition, we describe the challenge structure, evaluation criteria, and the winning developed models. Moreover, we detail novel interpretations of feature influences on the re-hospitalisation of Croatian cardiology patients. Finally, we assess the impacts of crowdsourcing competitions in the digital healthcare, in particular with respect to EDIH goals and impacts in the overall digital innovation ecosystem building and implicit stakeholder learning-through-doing.

## Materials and methods

Competition was organised by the AI4Health.Cro EDIH Consortium, with the aim to introduce crowdsourcing challenge as a tool to build awareness of AI capabilities, bring together young technology and entrepreneurial talent and initialise prototype solutions for the applications in real-life setting. Organisational team included a group of multi-disciplinary experts in medicine, public health, health technology assessment, AI and digital health research, with previous experience in hackathon organisation. The prediction of risk or re-hospitalisation was selected based on its overall and local relevance, taking into account data availability within the hospital information system (HIS), as well as evidence and recommendations from current medical literature. Besides the development of the diagnostic model evaluated on its predictive performance, the focus was on trustworthiness of the solution, i.e. interpretability of the generated model, particularly on the level of individual decisions. Of equal importance, aspects related to the usability and overall healthcare utility of the model and business aspects of the solution were additionally evaluated.

### Setting up the infrastructure

Challenge organisation required piloting processes related to acquisition of data from local hospitals, into datasets targeting research or innovation problem for secondary data use. This included obtaining ethical approval from medical institution, preprocessing, anonymisation and documenting the data use. In addition a SPE was provided to each participating team in phase 2 (Fig 1), on a virtual machine with 16GB of RAM, 4 CPUs, and 2 SSDs with 40GB for the OS and 100GB for data. The virtual machines were restricted to local operations, with internet access and connections outside the virtual machine blocked by firewall. The only allowed connections were from a virtual private network and to an secure file transfer protocol server, located on the same hardware as the SPE, for secure submission of proposed task solutions for evaluation.

**Fig 1.**
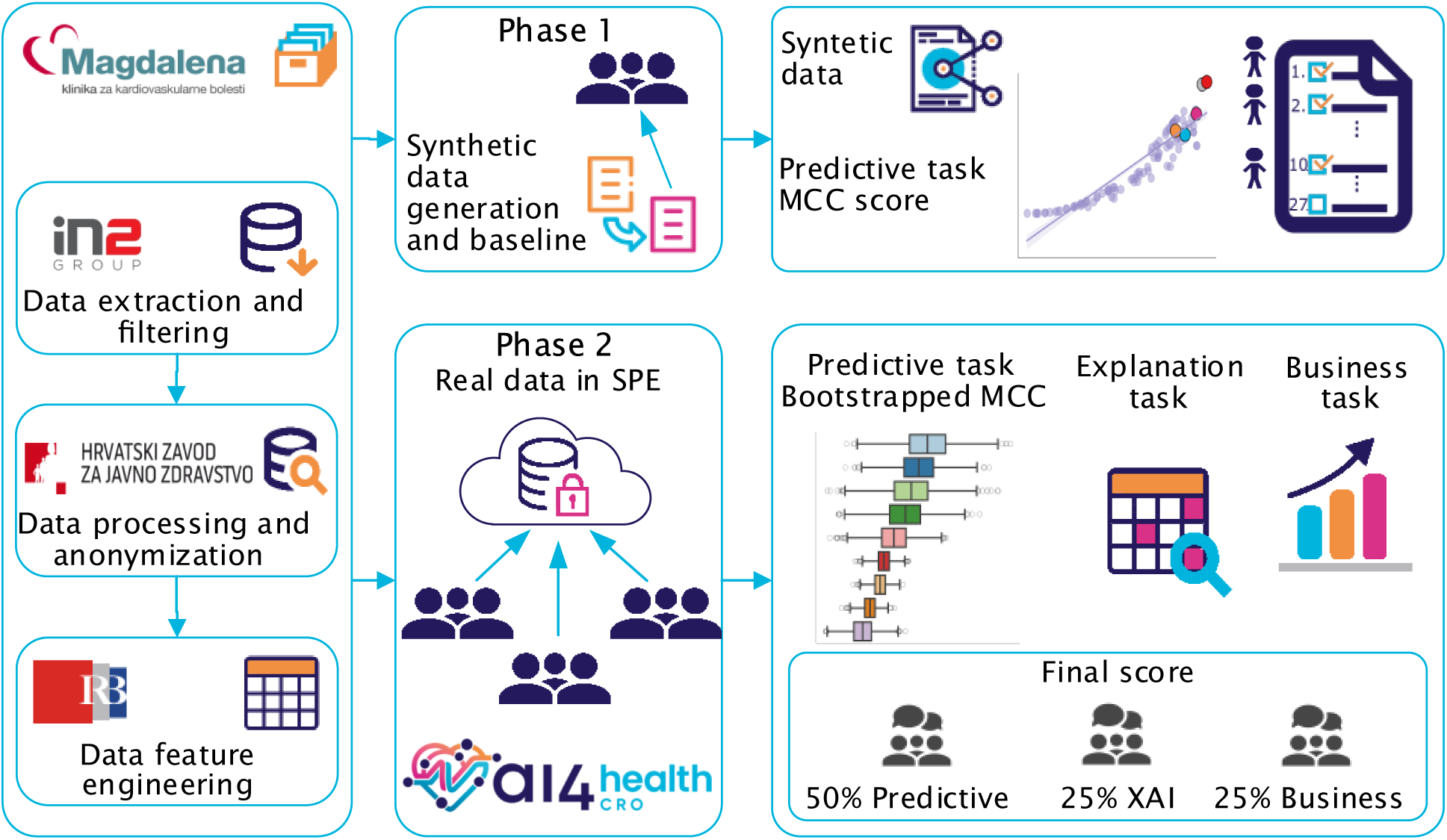
Re-hospitalisation competition workflow. Schematic overview of the re-hospitalisation competition workflow, from feature selection, data extraction, and anonymisation, to the creation of a synthetic dataset for phase 1, followed by phase 2 in which participants accessed the real data within a secure environment, and the subsequent evaluation.

### Challenge organisation and structure

Crowdsourcing challenge started with the pre-selection of teams of 2-5 participants, set in eligibility rules, often resulting in multidisciplinary teams [17]. Due to a large number of participants, competition was organised in two phases (Fig 1). Phase 1 was elimination round based solely on predictive performance of the machine learning model on a synthetic dataset, organised to filter top teams that were, in the second phase, admitted to SPE on Croatian Institute of Public Health (CIPH) servers. A total of 98 participants applied to the Challenge, forming 28 teams, with top 10 teams advancing to the second phase. Participants who qualified for the second phase of challenge were required to sign a Non-Disclosure Agreement before getting access to anonymised real-world data.

The first phase evaluation process involved daily solution submissions, with leaderboard containing a set of predictive performance metrics: Matthews correlation coefficient (MCC) [18], h-measure [19], F1, area under receiver operating curve (ROC AUC) and accuracy scores. The selection of top 10 performing solutions (teams) to continue in the second phase was based on the MCC score. They were then introduced to SPE and challenge rules for the second phase. Communication with competition participants primarily occurred through the competition forum, with two live meetup events at the beginning of the two phases. During the six weeks of the second phase of the competition, competitors were allowed to submit maximum of six prediction solutions. The final score of predictive models was based on teams last submission.

The overall evaluation of participant solutions consisted of three separate tasks: (i) the predictive task of quantifying the risk of re-hospitalisation; (ii) medical explainability about understanding predictions and explaining them to the target users, i.e. medical practitioners; (iii) business model and application prototype of the user interface that would be used in a hospital setting.

### Data acquisition and preprocessing

Data Processing Agreement accompanied by a data handling protocol were established between CIPH and Magdalena Clinic prior to data access. The agreement defined the legal basis, scope, and conditions for processing health-related personal data for research and innovation purposes. The protocol specified the categories of data processed, the purpose and duration of processing, the roles and responsibilities of participating institutions, and the technical and organisational measures used to ensure data security. The IT service provider IN2, responsible for the HIS at Magdalena Clinic, performed data extraction and CIPH carried out the processing of personal data in accordance to the protocol and GDPR provisions applicable to tasks carried out in the public interest and legitimate research activities. Given the involvement of sensitive health data, additional safeguards were implemented: data minimisation, controlled access, and protection of the rights of data subjects.

Extracted data records spanned the years 2014–2023 (Fig 2). Records from the COVID-19 pandemic years (2020, 2021, and 2022) were excluded, resulting in two disjoint subsets: 2014–2019 and 2023. The 2023 data differed substantially from earlier years due to the impact of COVID-19 on population health status, as well as changes in clinical practices. As a result, the 2023 subset was treated as an out-of-distribution (OOD) dataset. In total, the dataset comprised 46903 records and 27 variables, (S1 Appendix), and 481973 individual medication/substance entries, with a binary outcome label indicating occurrence of readmission.

**Fig 2.**
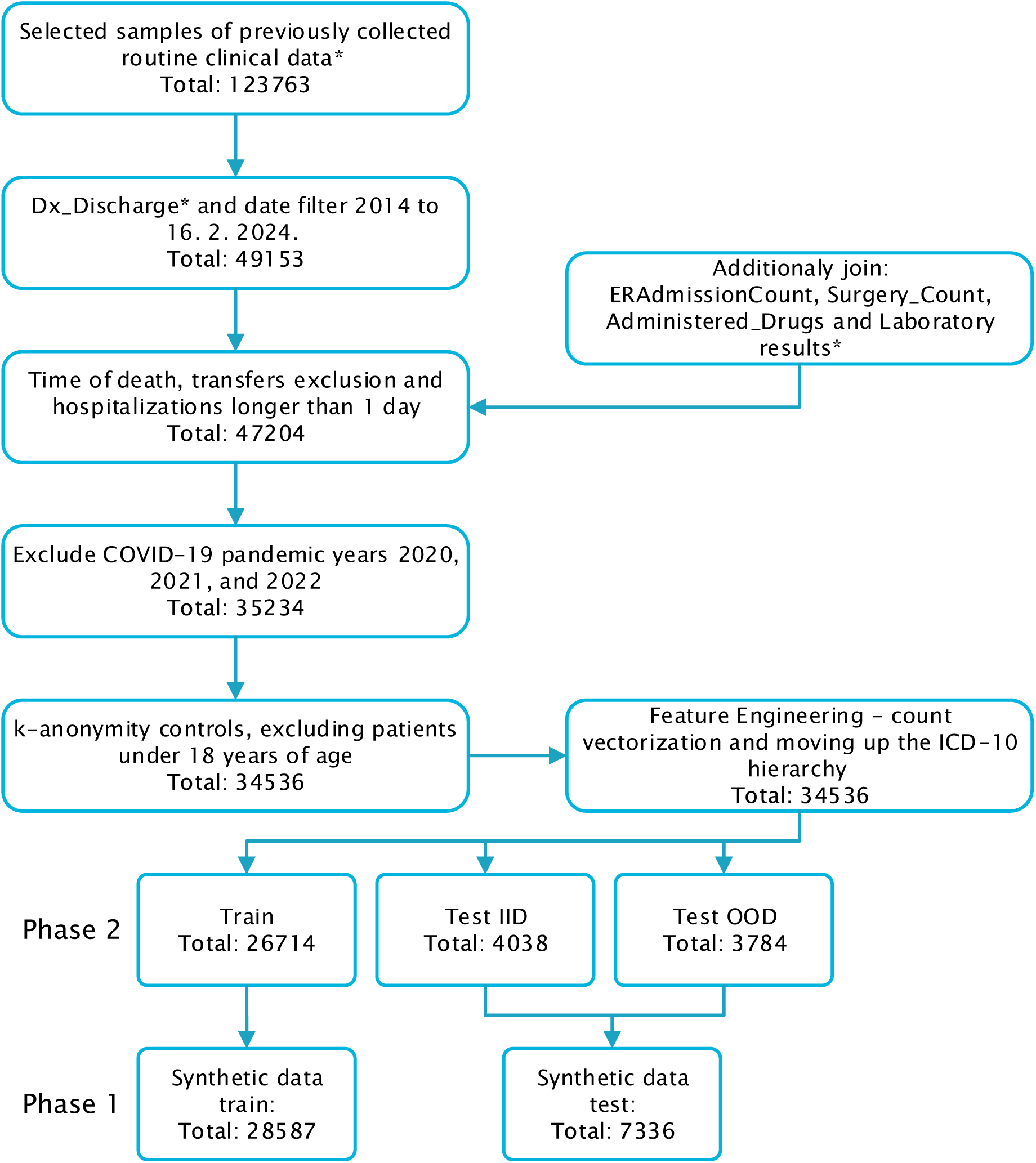
Data filtering and anonymisation workflow. Overview of the dataset reduction process from the initial 123763 samples to 34536 after filtering. The filtered dataset was split into train and test sets from which the synthetic train and test sets were generated, respectively. All intermediate steps and corresponding sample counts are shown in the figure. **\***Descriptions of each attribute are provided in S1 Appendix and the differences between original and synthetic dataset in S2 Appendix.

#### Anonymisation and processing

The data anonymisation process was carried out to protect patient privacy and ensure data security, in turn reducing the risk of misuse and re-identification of patients involved. Anonymisation procedures were applied to the dataset and enhancement was achieved through additional feature engineering, at the same time maintaining trade-off between data utility and privacy. All personal identifier variables were omitted.

Variables like “AdmissionDate” and “DischargeDate” were used in process of creating labels and were afterwards omitted. The “Age_Discharge” variable was processed by excluding all subjects under 18 years old. Additionally, a new variable was derived, binning patients into five-year age groups starting from 18 years old.

The 10^th^ revision of the International Statistical Classification of Diseases and Related Health Problems by the World Health Organisation, hereafter referred to as the ICD−10 [20] was used for disease coding. To improve k-anonymity and enhance data privacy, we transformed the ICD−10 codes for both admission and discharge diagnoses by grouping the hierarchical tree of the ICD−10 codes to the level of the initial letter, representing broader diagnostic categories. For example, the code I23.2 was generalised to I, Fig 3 a) and b). Secondary diagnosis contained additional conditions or complications a patient has alongside the primary diagnosis, including co-morbidities, conditions discovered during hospital stays, and any complications that arose, in the form of the ICD−10 codes. Count vectorisation was applied to the secondary diagnosis data. After simplifying each ICD−10 code to its initial letter, 25 features were created for each sample, where each feature corresponds to one of these ICD−10 categories. This transformation counted the occurrences of each ICD−10 category letter within the secondary diagnosis list and populated the feature values accordingly.

**Fig 3.**
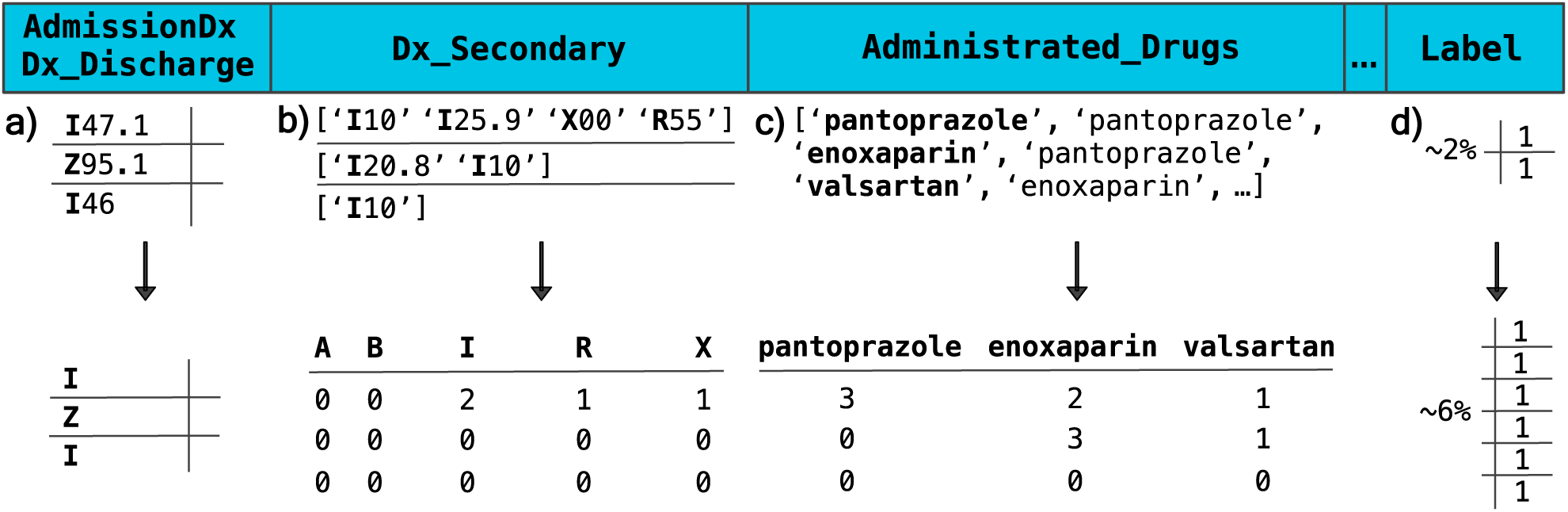
Feature engineering for synthetic data generation. Transformation of sparse clinical variables into higher-level representations. **a)** “AdmissionDX” and “Dx_Discharge” were encoded into categories based on the first letter of ICD−10 codes. **b,c)** The categorical lists “Dx_Secondary” and “Drugs_Administered” were vectorised using count encoding. For “Dx_Secondary’, ICD−10 codes were grouped at the chapter level, and occurrences were counted. For “Drugs_Administered’, entries were split into individual drug variables based on unique values, with counts reflecting repeated administration. After feature engineering, variables were filtered to meet sparsity constraints of the synthetic data generator, resulting in 21 ICD-10-based variables and 237 drug-related variables. **d)** Synthetic data were used to mitigate class imbalance in the target variable “Label’, increasing the minority class proportion from 2.296% to 6.61%.

A separate variable containing a list of drug codes was defined that represented therapy administered to a patient. In this context, anatomical therapeutic chemical (ATC) version 10 codes were converted to ATC−7 codes, eliminating the quantity, duration, and method of drug administration. Subsequently, ATC−7 codes were mapped to the corresponding generic names of the drugs. Finally, the “Drug_Administered” attribute was transformed similarly to the secondary diagnosis using count vectorisation, Fig 3. This process resulted in 303 additional features that captured the frequency of each administered drug category. After preprocessing and anonymisation, our data contained 34536 records and 347 variables. Differences between the original and synthetic data variables are detailed in (S2 Appendix).

#### Synthetic dataset

Synthetic re-hospitalisation dataset was generated to enable broad access to healthcare data (S1 Data), and experience of predictive modelling to all applicants of the competition, as SPE resources were too limited to accommodate all teams bridging the data access gap [21]. Synthetic data was generated using synthpop R platform package [22], and resulted in 28587 training set samples. Aggregation of features through feature engineering as shown in Fig 3 reduced sparsity over the domain of ICD−10 codes, while preserving other data properties. Utility of synthetic data was evaluated using propensity score mean squared error (pMSE) [23], Fig 4.

**Fig 4.**
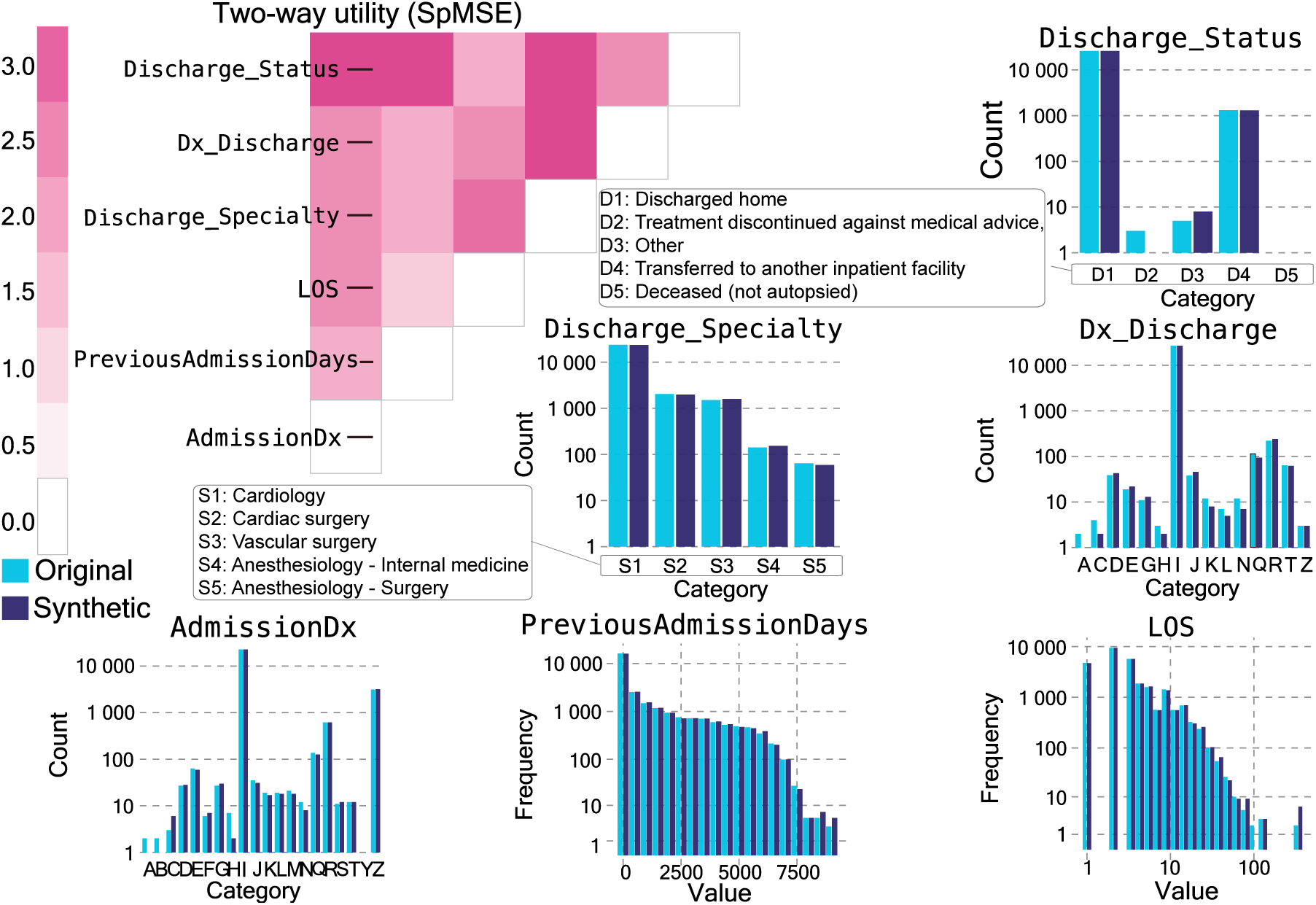
Comparison of synthetic (light blue) and original (dark blue) variable distributions. Histogram values are log-transformed. Variables were selected based on the highest SHAP values [12] in contestants reported explanations. The heatmap shows distances between propensity score distributions of synthetic and original features. Low standardised pSME values (e.g., “AdmissionDx’, “PreviousAdmissionDays’, length-of-stay “LOS”) indicate that the synthetic data retains the statistical characteristics of the original data. High values (e.g., “Discharge_Specialty’, “Discharge_Status’, “Dx_Discharge”) imply poorer performance in replicating specific patterns and relationships found in the original data, but enhance privacy making it harder to match back to real patients [24]. Translations of categorical values can be found in (S1 Table)

During generation process, we increased the prevalence of rehospitalized patients from 2.296% within the original data to 6.61% in the synthetic training data. For the generated synthetic test set of 7336 samples, the prevalence of positive class was 5.65%. In this way, the true distribution of positive class remained unknown to the participants of the challenge, preventing information leaks (S1 Code).

#### Anonymised re-hospitalisation dataset

The anonymised real re-hospitalisation data has been used in the second, restricted phase of the challenge. The training set contains all patients up to and including year 2018 and 20% of samples stratified on binary feature one and more than one hospitalisation from the year 2019. The test set contains rest of the samples from the year 2019 and all the samples in the year 2023. We emphasise the high imbalance in the resulting training and validation datasets with only 2.296% readmitted patient, which is significantly diverging from the average of 11.95% reported by previous re-hospitalisation studies.

Since the participants were working within the SPE, the second phase dataset retained more of its original form compared to the synthetic dataset. The true dataset included an additional variable, “AdmissionYear’, which indicated the year the patient was admitted to the hospital. Admission and discharge diagnoses, “Administered Drugs” and “Secondary Diagnoses” were preserved in their original structure, maintaining the full structure of ICD−10 code.

### Tasks and evaluation of solutions

Phase 1 of the challenge was evaluated solely by MCC on a synthetic dataset. Phase 2 evaluation score had three tasks: predictive (50 points), medical explainability (25 points), and business solution (25 points). Evaluation of final phase two solutions was performed by nine evaluators: three technical, three medical and three business evaluators. Each group assessed solutions using a 0 − 5 point scale for each of the evaluation criteria listed in the Table 1. Final score that determined the winning solution was the sum of three tasks.

**Table 1.** Technical, medical and business evaluation criteria.

| **Technical evaluation** |  |
| --- | --- |
| Checklist | Submitted (yes/no) |
| Technical documentation |  |
| Source code |  |
| Model predictions |  |
| Medical documentation |  |
| Notebook for medical findings |  |
| Prototype solution link |  |
| Video pitch link |  |
| Criteria for medical explainability | 0-5 point scale |
| Analytical capabilities identifying the most important variables |  |
| Analysing their interactions |  |
| Discovering and understanding patient subgroups particularly those with an increased likelihood of early rehospitalisation |  |
| Identifying biases in the data or the developed model |  |
| Overall coding clarity |  |
| Total technical evaluation of medical explainability | **TEC** |
| **Medical evaluation** |  |
| Criteria for medical explainability | 0-5 point scale |
| Innovation |  |
| Health outcomes |  |
| Patient relationships |  |
| Compliance |  |
| Ethics |  |
| Total medical evaluation of medical explainability | **MED** |
| **Total evaluation of medical explainability** | **Mean(TEC, MED)** |
| Criteria for business model proposals | 0-5 point scale |
| Innovation |  |
| Health outcomes |  |
| Patient relationships |  |
| Compliance |  |
| Ethics |  |
| Total medical evaluation of business model proposals | **MBUS** |
| **Business evaluation** |  |
| Criteria for business task | 0-5 point scale |
| Innovation |  |
| Market potential |  |
| Team quality |  |
| Sustainability |  |
| Scalability |  |
| Total business evaluation of business model proposals | **BUS** |
| **Total evaluation of business model proposals** | **Mean(MBUS, BUS)** |

For solution to be eligible the existence of all scripts for reproducibility of the model and documentation was mandatory or the solution would not be eligible for evaluation. Total technical (TEC), or medical (MED), or medical business (MBUS) or business (BUS) evaluation of a task was an average between the three evaluators.

#### Predictive task

Test set included the independently identically distributed data (IID) corresponding to samples between years 2014 to 2019 and out-of-distribution (OOD) data corresponding to year 2023. Teams final score was produced by bootstrapping the test set 2000 times and taking the mean value of the weighted MCC scores, the 0.75*∗*MCC_IID and 0.25*∗*MCC_OOD, on each of the bootstrap samples, *i*:

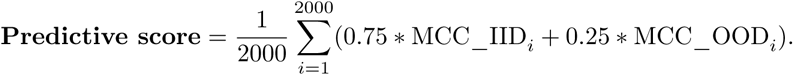

Firstly, teams were sorted in descending order by the calculated **Predictive score**. Secondly, the maximum number of points was given to the first team. Then all other teams were awarded an aliquot fewer points received based on percent of difference to the immediate predecessor in final predictive score e.g. here team A has the highest final score *x* followed by Team B the second highest final score *y*:

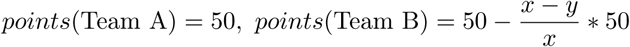

#### Medical explainability

The prototypes had to be developed with clinical staff, nurses and physicians as the target end-users of the predictive models. Accordingly, participants were expected to explain the re-hospitalisation risk of individual patients, contextualising predictions and highlighting key data interactions indicative of severe clinical states.

Participants were asked to submit both a textual report and accompanying script or notebook demonstrating how they derived their conclusions from the data. The goal was to assess the ability to generate clinically relevant and interpretable explanations for healthcare professionals.

The report had to: a) identify the most important variables contributing to the prediction, b) analyse interactions between these variables, c) discover and characterise patient subgroups, with an emphasis on those at higher risk for early re-hospitalisation, d) detect biases either present in the dataset or introduced by the developed model, e) communicate findings in a format that is accessible and informative to clinical stakeholders. Technical judges and medical judges evaluated the teams submitted results based on criteria in Table 1.

#### Business model proposals

The Business Task required participants to develop a prototype application interface designed for use in a hospital setting [2]. The application was expected to: a) provide risk level assessments for individual patient cases, b) be supported by explanations of key contributing factors, c) leverage user experience, and d) provide an assessment of prediction uncertainty.

In addition to the prototype, participants were asked to prepare a video pitch of up to 3 minutes, highlighting the aspects like potential value, feasibility, and implementation strategy. The business judges criteria are in Table 1.

## Results

The implementation of the AI4Health.Cro Challenge demonstrated a successful crowdsourcing experiment across participating institutions and teams. A total of 27 teams engaged in the challenge and in phase 2 seven teams out of 10 completed all tasks. Engagement was sustained across both phases of the competition, including a transition from synthetic data to real-world clinical data, indicating adaptability of participants to increasing data governance requirements. Engagement of teams with the organisers via an online discussion forum resulted in 27 topics and 107 posts over the course of the competition, reflecting the complexity of proposed multidisciplinary problem. A total of 115 valid submissions were received in phase 1 (Fig 5), followed by 40 submissions in phase 2 (Fig 6). In phase 1 best performing classifiers exhibit similar ROC AUC score (Fig 5e). The mutual information (MI) estimation approximated with k nearest neighbours is between MCC and H-measure 1.74, with Spearman-rank correlation of 0.91, Fig 5c.

**Fig 5.**
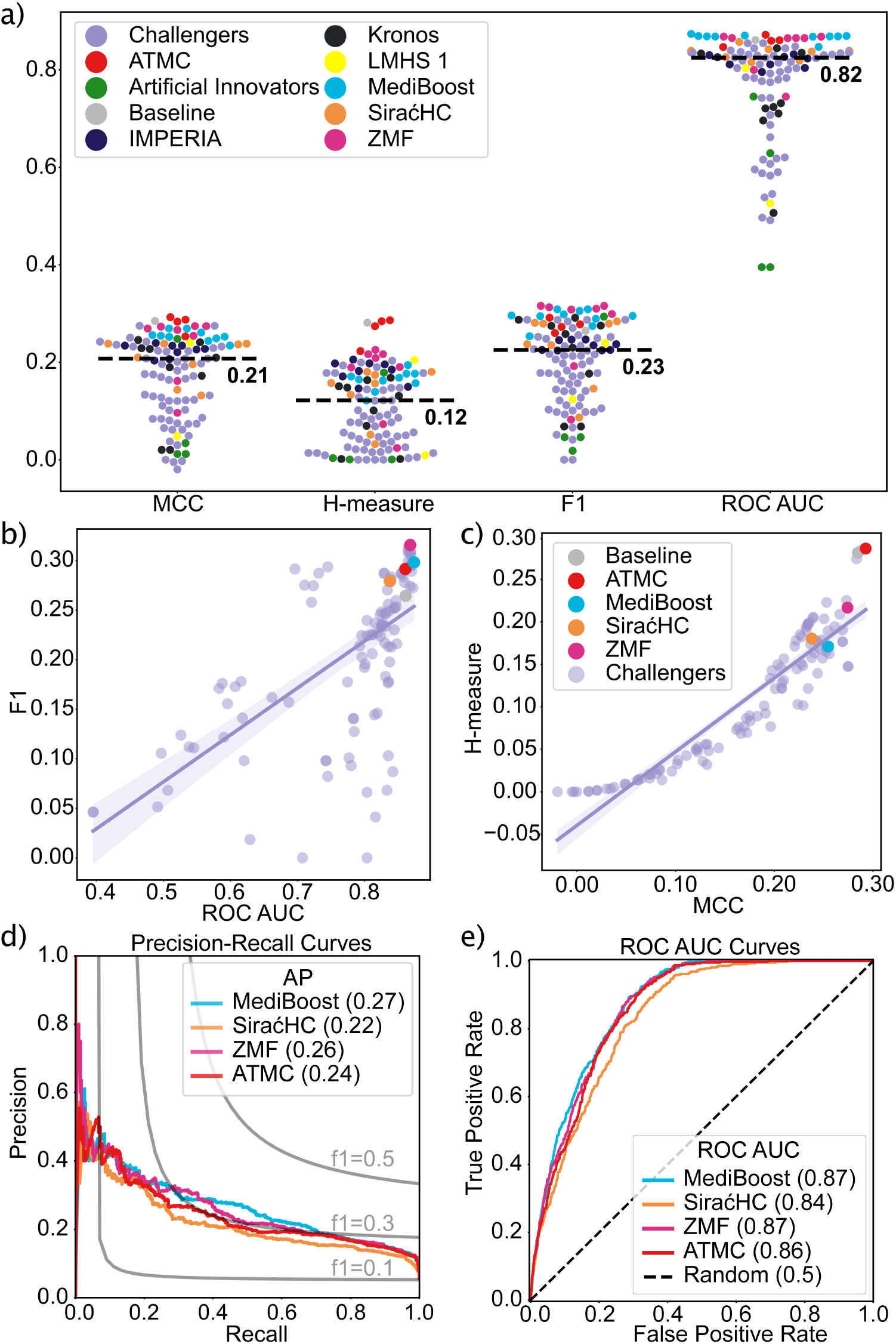
Performance of teams on synthetic data in phase 1. Image **a)** describes the contestant submissions with respect to MCC, H-measure [19], F1 score and ROC AUC. Purple colour represents submissions of all other teams, labelled as Challengers. Images **b)**, **c)**, **d)** and **e)** legend represents the phase 1 winning team final submission and top three teams, winners, from phase 2, and their last submission in phase 1. Both images **b)** and **c)** show scatter plot of ROC AUC and F1 score, and MCC and H-measure. Compared to MI estimation between F1 - MCC = 1.39, and ROC AUC - MCC = 0.79. **d)** Precision-recall curves are generated from final submissions probabilities of class readmission on the test set. **e)** Test set evaluation of best performing teams ROC AUC are within 3.5% range. For any of these classifiers we could use lift at threshold 0.9 to identify more than 40% of potential patients that will be readmitted within 30 days and analyse the preventive methods and procedures that this risky group should undergo.

**Fig 6.**
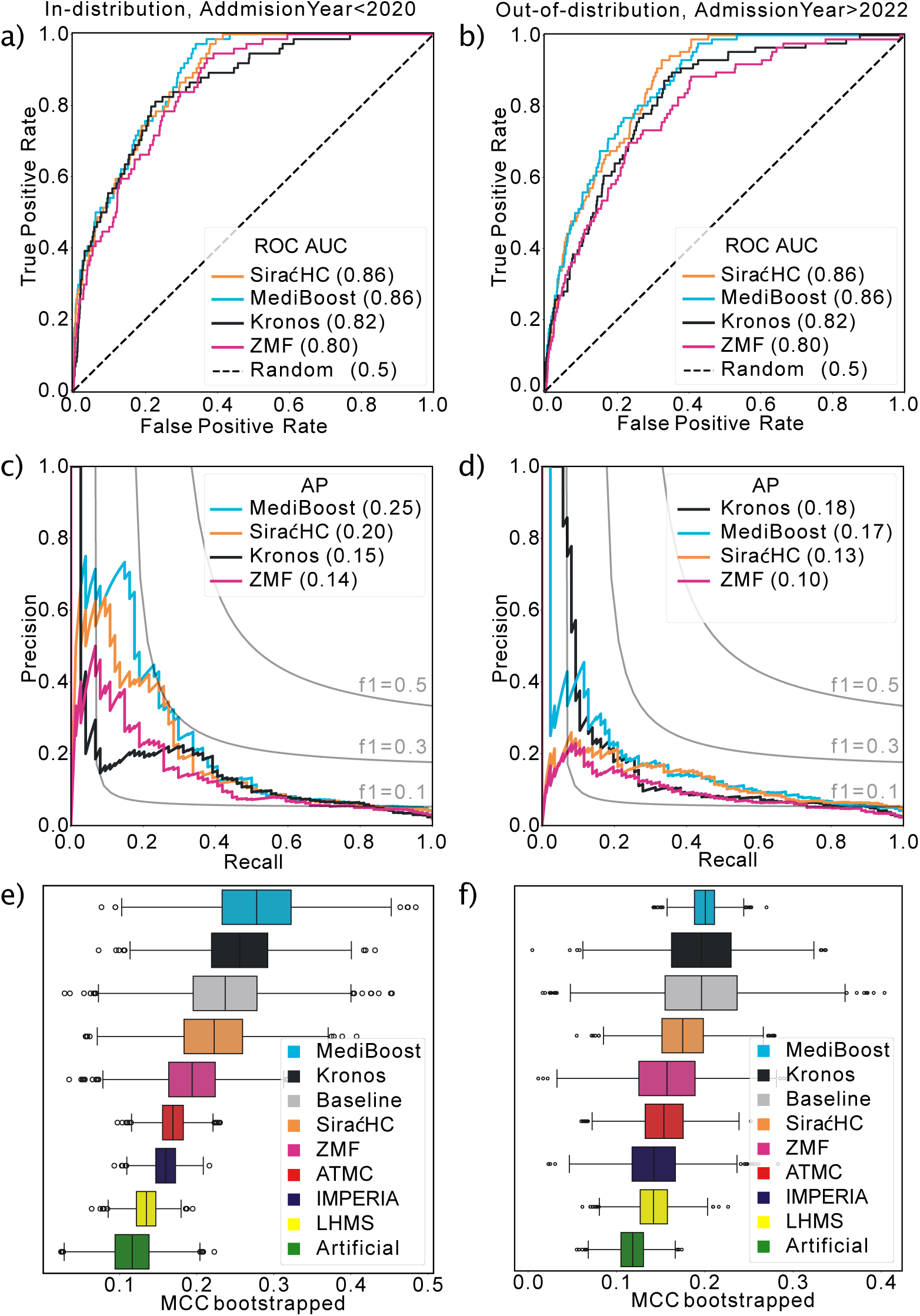
Phase 2 IID and OOD predictions. Left column of images are results on the in-distribution data data gathered between 2013 − 2019. The right column of images is the OOD test data evaluation. **a)**, **b)**, **c)** and **d)** images contain the final submission made in secure processing environment (SPE) by top 4 teams in the challenge. In **a)** we can identify two highest AUC scores from MediBoost and SiraćHC teams. Recall and precision on **c)** and **d)** graphs are low due to highly unbalanced data. **e)** and **f)** are the results of 2000 bootstraps on the test set for predictions of in-distribution and OOD data. Most solutions fall within each others 95% confidence interval.

From an organisational perspective, the availability of a centrally managed data governance framework and SPE supported effective execution of the challenge. All teams conducted their analyses within restricted virtual machines, submitted solutions exclusively via the designated secure file transfer mechanism, and complied with predefined access limitations. No protocol deviations, compliance-related interruptions, or data security incidents were observed during the competition period.

Governance responsibilities related to data access and security were managed centrally by the organising institutions, allowing participants to focus on methodological development while adhering to predefined legal and technical requirements. Teams predominantly adopted ensemble-based machine learning approaches and optimised model performance according to the predefined evaluation criteria. The submitted models’ characteristics reflect both the constraints and affordances of the implementation setting, including requirements for transparency, explainability, and secure execution.

### Developed solutions

We hereby present the top three solutions, scored according to the procedure detailed in Section Tasks and evaluation of solutions. Each competitors choice of key feature engineering strategies and the underlying modelling techniques is shown below. To contextualise these results, Fig 6 illustrates both in-distribution and OOD evaluation outcomes, highlighting variability in generalisation and the impact of dataset shift across submissions. Overall, while the top-performing models achieve comparable discrimination as reflected by overlapping confidence intervals, their differing characteristics reveal important trade-offs between robustness, precision, and sensitivity for this highly imbalanced task.

#### MediBoost

Team Mediboost’s methodology encompassed comprehensive data processing, predictive modelling, and explainability analysis. During data cleaning, missing values were handled based on variable type: continuous variables were imputed with median values, categorical variables were explicitly labelled as missing, and missing instances of “PreviousAdmissionDays” were assigned a constant value of 9000. Extreme outliers in height and weight were replaced with median values, accompanied by a new indicator column for missing-ness. Logical corrections were also applied to instances where height and weight were erroneously swapped. To approximate a normal distribution, the “LOS” was log-transformed.

Categorical variables underwent strategic grouping; referral and discharge diagnoses were aggregated into ICD-10 subgroups, preserving crucial three-letter codes, while Z diagnoses were grouped separately.

Demographic data were streamlined by grouping educational status into low, medium, and high tiers, and employment status into employed, inactive, and retired categories. Furthermore, rare clinic codes were consolidated, and age categories were converted into a continuous variable. Feature engineering introduced a binary indicator for previous hospitalisations within the past month, alongside specific flags for diabetes, chronic obstructive pulmonary disease (COPD), chronic kidney disease, and hypercholesterolaemia. Cardiovascular diagnoses were consolidated into binary features. Medication features were filtered to retain only those present in at least 3% of cases, and new variables were created to capture the total volume and variety of administered drugs. The dataset was partitioned using three-fold cross-validation, stratified by target class and admission year.

For modelling, a three-layer neural network (with 512, 256, and 128 neurons) generated representation vectors for categorical features. The network was optimised via Bayesian methods and trained for 15 epochs using a variable learning rate, AdamW optimisation, L2 regularisation, and a 0.6 dropout rate. These learned embeddings, integrated with numerical features, served as inputs for XGBoost and CatBoost algorithms. Both tree-based models were similarly optimised using Bayesian optimisation and cross-validation. The final architecture linearly combined these models across each cross-validation fold, yielding a robust six-model ensemble.

#### SiraćHC

The dataset underwent preprocessing to ensure data integrity and accuracy, including fixing implausible values. Weights exceeding 1000 were assumed to be missing a decimal point and were divided by 100.

Weights over 210 were divided by 10, and weights under 40 were converted to missing values. Similarly, for heights, values below 100 were increased by 100, and any implausible values that could not be reasonably corrected were converted to missing values. Additionally, employment status was converted to a factor with unknown value changed to missing, age groups were turned into a numeric variable representing the average age from the range. Diagnosis values were truncated to their first letter. Redundant variables such as “Secondary Diagnoses’, “AdmissionYear” and “AdministeredDrugs” were removed. Body Mass Index (BMI) was calculated from height and weight.

A 3-fold cross-validation with a proportional data split was conducted, using ROC AUC as the performance metric. The team employed the tree-based XGBoost algorithm due to its ability to handle missing data without requiring imputation and accommodate diverse data types, which was crucial given the nature of the data. For model training, XGBoost and caret packages in R were used to select optimal hyperparameters. A weight of 10 for positive prediction class.

#### ZMF

Data cleaning and feature engineering included: converting “Discharge_Specialty” to string; imputing missing “PreviousAdmissionDays’, weight, and height with mean/median and adding missing indicators; replacing missing Education and “Current_Work_Status” with unknown; and one-hot encoding categorical variables. “AdmissionYear” was removed, but pre-COVID (2019 and before) and unknown year indicators were added.

In terms of feature construction, newly constructed variables significantly enhanced prediction: Total drug count, 30 drug groups amounts, calculated BMI, total diagnosis count, and a length-of-stay “LOS” multiplied by total drug count variable. A simplified CCI was included, along with binary indicators for CCI*>* 6 and unknown CCI. Removing “Administered Drugs” and “Secondary diagnoses” resulted in 528 total columns.

The model was developed using original and synthetic data. A test set (10% of original data) was stratified, and the remaining 90% was used for 5−fold stratified cross-validation to select the best model (logistic regression, decision trees, XGBoost). Logistic regression was chosen for its interpretability and efficiency, ultimately performing best after training on combined data with optimised hyperparameters. Hyperparameter optimisation (grid and random search) maximised the average MCC across five folds. The final model was retrained on the full training set and evaluated on the test set using MCC, precision, recall, and F1 score. An optimal probability threshold (typically 0.5 − 0.7) was then found to balance precision and recall. To enhance positive-class recall due to strong dataset imbalance, misclassification of positive class was heavily penalised, resulting in more positive examples detected. Model explainability is achieved via variable selection and logistic regression. Variables were selected based on Chi-squared measure for linear dependence with the outcome, reducing initial 528 variables to 300.

### Model explainability

Teams had to produce an explanation of their model predictions and findings for the medical evaluation. Evaluation and scoring was done independently by 3 evaluators that based their sores on 5 criteria using 25-point scale for detailed description of evaluation, scoring, instructions, and medical explanation task in general see Section Medical explainability. In the following sections we provide a brief summary of competing teams explanations.

#### MediBoost

Feature importance analysis identified “PreviousAdmissionDays” as the strongest predictor, followed closely by discharge diagnoses (“Dx_Discharge”), which critically differentiated between chronic and life-threatening conditions. “AdmissionYear” also demonstrated high predictive value. Among specific conditions, Z diagnoses particularly those indicating cardiac and vascular implants (Z95) were highly influential. Crucial medication predictors included overall drug volume, variety, and the use of iohexol. Conversely, variables like “LOS_ICU” provided negligible signal.

SHAP further elucidated model behaviour, revealing that longer intervals since the previous hospitalisation correlated with higher re-hospitalisation risk. Notably, the 9000 placeholder substantially increased predicted risk, likely because an absence of hospital records suggests irregular care. Higher risks were also associated with non-home discharges, prolonged “LOS’, and multiple cardiovascular co-morbidities. In contrast, risk estimates decreased for patients with lower discharge weights, a higher number of past surgeries, Z diagnoses, and later admission years. Finally, specific medications driving predictions included enoxaparin, warfarin, allopurinol, and nitrazepam.

#### SiraćHC

In the context of XGBoost models, understanding the metrics of gain, cover, and frequency is crucial for interpreting and explaining the model’s behaviour and performance. The gain metric which reflected the improvement in model accuracy through variable splits, showed that “PreviousAdmissionDays’, “Discharge Status”: other healthcare facility, and ICD−10 code I cardiovascular disease, were particularly influential. These variables significantly enhanced the model’s predictive accuracy. The frequency metric, which measured how often variables were used in tree splits, highlighted that “PreviousAdmissionDays’, ICD−10 chapter I, and “Discharge Speciality”: cardiac surgery were frequently employed. Among the most important features was the ICD−10 chapter I, which encompassed various chronic conditions and complex medical diagnoses that heighten the risk of early re-hospitalisation. This code captured conditions such as heart failure and COPD, which are known to exacerbate and lead to frequent readmissions. BMI also emerged as a significant variable. Both obesity and underweight can affect health outcomes and increase the risk of complications, contributing to re-hospitalisation. Treatment with ioversol, a contrast agent used in imaging, was another critical feature due to its association with potential side effects and complications leading to re-hospitalisation. Similarly, the use of warfarin, an anticoagulant, was pivotal due to its impact on conditions like pulmonary embolism and thrombosis, which could result in readmission if not managed properly. Additional important features included the “LOS’, which often indicated the severity of the condition or complications encountered.

#### ZMF

The logistic regression model threshold was adjusted experimentally to 0.642 to optimise performance. The model uses the log-odds derived from its parameters and input variables.

Each variables parameter value shows its influence on re-hospitalisation likelihood; a higher absolute value means a greater impact on log-odds and predicted probability. Key considerations are that coefficients, determined post-training, might vary in importance on the test set, and unscaled numerical variables may misrepresent their true impact, especially those with large ranges. Positive parameters substantially increase re-hospitalisation odds, while negative ones decrease them, directly indicating the direction of influence since all training values were non-negative. This methodology ensures clear interpretability of each variables predictive impact.

The top three categorical variables associated with increased re-hospitalisation risk are: “PreviousAdmissionDays” - unknown, “Discharge_Dx” - N (urogenital), and “AdmissionDx” - unknown. Factors decreasing risk include: “Discharge Status” - Home, “AdmissionDx” - G (central nervous system), and “AdmissionDx” - M (musculoskeletal).

Numerical variables associated with increased re-hospitalisation risk are: glucose count, salmeterol count, and ampicillin/betalactamase inhibitor. Variables associated with reduced re-hospitalisation risk are: levobupivacaine, lisinopril, and ceftazidime. Marital status and education showed no significant linear dependencies, suggesting no bias.

## Discussion

### Main aims

Our aim in organising the crowdsourcing Challenge in digital healthcare realm was to proactively boost the healthcare innovation ecosystem, which is one of the key major objectives of EDIH. Crowdsourcing approach impacted mainly future talent in both, AI and healthcare domain, but also institutional players directly related to aims of digital transformation of both, small innovative companies and medical domain stakeholders. Other aims included: raising awareness on the role of AI in healthcare as well as on complexities related to developing applications based on primary medical data; piloting the process of secondary data use according to new EU regulations; testing complexity and potential utility of new AI based tool in the context of the relevant problem in healthcare.

### Impacts

In the specific case of the healthcare domain required collaboration between clinical, technical, and research stakeholders within the AI4Health.Cro EDIH to organise a challenge and provide sandbox-like environment for data driven crowdsourcing research endeavour. The establishment of a formal data processing agreement and accompanying protocol played a central role in enabling the implementation of the AI4Health.Cro Challenge. By clarifying legal responsibilities, data access conditions, and safeguards prior to analysis, the governance framework reduced institutional uncertainty [2], and facilitated collaboration between clinical, technical, and research stakeholders [4]. This structure allowed public participants to focus on methodological innovation and problem solving rather than on ad-hoc compliance considerations, highlighting the importance of legal and organisational preparedness and multi-institutional involvement in healthcare secondary data use. With respect to healthcare digital innovation ecosystem the challenge elicited forming of informal groups of young AI and medical talents on collaborative complex problem solving. Lasting almost two months, this effort resulted with implicit learning experience for the involved participants, demonstrating in practice what is required in order to innovate in digital healthcare: the structured process of data governance, provision of secure processing environment by the AI4Health.Cro and specific requirements and evaluation to construct a trustworthy AI application through the crowdsourcing effort.

### Participants and solutions

Teams applying to the challenge that included medical experts scored better and described/explained their models and solutions (e.g. feature importances, patient segments) significantly better than other teams, aligning their solutions very well with the medical knowledge regarding cardiovascular patients. Tree based models had high overlap in important features while logistic regression had different important features and ability to discriminate between positive and negative influence to the model decision. One of the (common and plausible) observations from the solutions is that certain medical conditions, such as heart failure, COPD, or oncological diseases, inherently carry a higher risk of re-hospitalisation due to the severity of the illness itself. Also, early readmissions often highlight challenges in care coordination, discharge planning, or inadequate patient support after leaving the hospital - which are findings from previous similar studies reported in literature. This issue is compelling because solutions developed through the analysis of early readmissions can benefit both hospital physicians during post-discharge care and the patients themselves. The evaluators of developed solutions were convinced that with some adaptation of top solutions of the challenge could reduce the rate of re-hospitalisations.

### Lessons learned

As Croatia and other European Union member states move toward implementation of the EHDS, standardised procedures for accessing and processing health data may lower administrative barriers and improve the scalability and reproducibility of similar initiatives. Our findings suggest that harmonised governance frameworks, combined with secure processing environments are likely to be critical for enabling cross-institutional and cross-border collaboration in applied health data science. Findings of the challenge, supported largely also by clinical practice [25], demonstrate that relying solely on digital technology, algorithms and experts in AI, for deployment of decision support systems in clinical setting is insufficient. The role of medical professionals is critical in interpreting requirements for such systems as well as their multidisciplinary evaluation and tailoring for personalised decisions and treatments. This is even more relevant for patients with complex conditions, which is often true for cardiovascular patients, where a personalised approach is often essential for successful treatment. The organizers experience with data acquisition, anonymisation, SPE development and the design of evaluation strategies highlighted the importance of coordinating data governance, technical infrastructure and evaluation when organising the challenge in digital healthcare. These activities share features with the regulatory sandbox approach set out in Chapter VI of the AIA [14], aimed at the development, testing and evaluation of AI solutions in a controlled environment under the supervision of competent stakeholders. Although the regulation was formally adopted after the completion of the challenge, the study infrastructure demonstrates how similar research and innovation challenges or datathons in healthcare, can support research and innovation operating within governance frameworks consistent with emerging regulation.

## Conclusion

This crowdsourcing innovation challenge is demonstrated in the context of developing AI solutions for targeted problems in healthcare. As a strategy to boost multi-stakeholder engagement and healthcare innovation ecosystem building, it illustrates how EDIH-supported initiatives [16] operate effectively within the EHDS framework to advance predictive healthcare. This activity, however, is somewhat beyond typical EDIH roles and services: it builds awareness, promotes multidisciplinary approach to creation of digital interventions in healthcare and creation of potential novel innovation startups. The challenge organisation was also a demonstration or piloting of the regulatory sandbox approach for developing innovative AI solutions based on real medical data, in accordance with new EU regulations, such as EHDS and AIA, related to secondary data usage in EU and deployment of AI systems in healthcare. Altogether, this endeavour represented an implicit learning-through-doing experience, both for the challenge participants and organisers. The tangible scientific outcomes of the crowdsourcing innovation challenge, however, are the innovative prototype solutions that were evaluated for their predictive performance, trustworthy aspects and business requirements, all important facets of advanced digital healthcare solutions, that are prerequisites for their successful uptake in clinical setting.

## Supporting information

Appendix 1 Extracted attributes

Table translations

Appendix 2 Data descriptors

## Data Availability

The synthetic dataset is available for the public use online at University of Zagreb, University Computing Centre, Digital Academic Archives and Repositories, FULIR data, URN:NBN: https://urn.nsk.hr/urn:nbn:hr:241:237658,
Synthetic Cohort for 30-Day Re-hospitalization Prediction.

## Supporting information

**S1 Appendix. Extracted attributes.pdf** Contains description of each attribute.

**S2 Appendix. Dataset features descriptors.pdf** Table that contains attribute descriptions after filtering, anonymisation and feature engineering.

**S1 Data. Synthetic Cohort for 30-Day Rehospitalization Prediction.** A public repository is available at: https://data.fulir.irb.hr/en/object/irb:904. The final release consists of a training set and a test set with 274 features including demographics, medications, and ICD-10 comorbidities. The dataset preserves key statistical characteristics of the original cohort while enabling safe data sharing and research on rehospitalization risk without exposing sensitive patient information.

**S1 Table. Translations of features in Croatian.** Translations of Croatian feature values within the dataset.

**S1 Code.** https://github.com/mlkr-rbi/challenge_rehospitalization_2024. Code repository for Crowdsourcing AI Solutions in early rehospitalisation as a pilot for healthcare regulatory sandbox.

## Abbreviations

EDIH: European digital innovation hubs
AI: Artificial intelligence
GDPR: General Data Protection Regulation
EHDS: European Health Data Space
NN: neural networks
CCI: Charlson Co-morbidity Index
AIA: AI Act Regulation (EU) 2024*/*1689
SPE: Secure processing environment
CIPH: Croatian Institute of Public Health
MDR: Medical Device Regulation
HTAR: Health Technology Assessment Regulation
HIS: Hospital information system
MCC: Matthews correlation coefficient
ROC AUC: Receiver operating curve area under the curve
OOD: Out-of-distribution
ATC: Anatomical therapeutic chemical
pMSE: Propensity score mean squared error
IID: Independently identically distributed
MI: Mutual information
COPD: Chronic obstructive pulmonary disease;

## Author contributions

Conceptualisation: A.B., A.J., M.Pi., M.K., M.B., S.K., T.S., T.L., Z.P.; data curation: M.B., M.Š.-B. N.Š., P.I., I.K., M.K., S.K., Z.P.; formal analysis: M.B., M.K., I.K.; funding acquisition: T.L., A.B., T.S.; investigation: A.B., T.S., M.K.; methodology: A.B., T.S.,M.K.; project administration: P.Z., M.Pr., Ž.B.; resources: M.B., P.I., M.K.; software: I.K., M.B., M.K., A.Š., J.K., K.P., L.T.-G., M.T.-G., I.B., R.Š., F.M., F.Ð., Z.A., M.Pa.; supervision: T.S., A.B.; team-Mediboost: A.Š., J.K., K.P.; team-siraćHC: L.T.-G., M.T.-G., I.B., R.Š., F.M.; team-ZMF: F.Ð., Z.A., M.Pa.; validation: M.K., T.Š.; visualisation: I.K., M.K., M.B.; writing - original draft: I.K., M.B., M.K., D.O.; writing - review and editing: M.K., A.B., T.S.;

## Acknowledgements

We acknowledge support from “Artificial intelligence for smart healthcare and medicine, AI4Health.Cro” (DIGITAL-2021-EDIH-01-101083735) project of European Digital Innovation Hubs. Collaborators of EDIH project cherish the work put in by dr. sc. Tomislav Lipić our colleague who helped envision, conceptualise and realise AI4Health.Cro project of European Digital Innovation Hubs.

