## Appendix 1 Extracted attributes for "Crowdsourcing AI solutions in early rehospitalisation as a pilot for healthcare regulatory sandbox"

### S1 Appendix Extracted attributes

**Table 1: Initial query variables for the Hospital information system.**

| | Columns | Type[1] | Description |
|---|---|---|---|
| 1 | `Patient` | Num | Unique patient identifier. |
| 2 | `Age_Discharge` | Num | Age at discharge, in years. |
| 3 | `Gender` | Cat | Patient gender (binary: M, F). |
| 4 | `Clinic_Code` | Cat | Category representing the clinic at discharge within the hospital (e.g., cardiology, ICU). |
| 5 | `AdmissionDate` | Date | Date of hospital admission. |
| 6 | `DischargeDate` | Date | Patient discharge date. |
| 7 | `Dx_Discharge` | Cat | Diagnosis at discharge, encoded using ICD-10 codes. |
| 8 | `Dx_Secondary` | CatL | List of patient comorbidities, encoded using ICD-10 codes. |
| 9 | `AdmissionDx` | Cat | Diagnosis at admission, encoded using ICD-10 codes. |
| 10 | `Admission_Id` | Num | Serial number used as an admission timestamp, from which the 30-day rehospitalization variable is derived. |
| 11 | `LOS` | Num | Length of stay, in days. |
| 12 | `PreviousAdmissionDays` | Num | Time since previous hospitalization, in days; used to create the label. |
| 13 | `AdmissionType` | Cat | Admission type: 'emergency' (typically transfers from other hospitals) or 'elective' (planned admissions, not necessarily pre-scheduled treatment procedures). |
| 14 | `ERAdmissionCount` | Num | Number of visits to the emergency department in the previous six months. |
| 15 | `Discharge_Status` | Cat | Discharge status (e.g., home, transfer to another clinic, ongoing treatment). |
| 16 | `Administered_Drugs (ATK 7)` | CatL | Therapeutic drugs during hospitalization are recorded and encoded in ATK 7 - WHO codes. |
| 17 | `Surgery_Count` | Num | Number of surgical procedures during the hospital stay. |
| 18 | `LOS_ICU` | Num | Number of days spent in the intensive care unit (ICU). After surgery, or emergency department. |
| 19 | `AdmissionYear` | Num | Year of admission of the patient to hospital treatment. |
| 20 | `Weigth_Discharge` | Num | Last recorded weight of the patient during the hospitalization. |
| 21 | `Height_Discharge` | Num | Last recorded height of the patient during the hospitalization. |
| 22 | `Education` | Cat | Socioeconomic level of education, e.g. VSS - higher education, SSS - secondary education, ... |
| 23 | `Current_Work_Status` | Cat | Work status: employed, unemployed, ... |
| 24 | `Glukose` | Num | Laboratory work sugar (mmol/l)[2] |
| 25 | `Potassium` | Num | Laboratory work potassium (mmol/l)[2] |
| 26 | `Sodium` | Num | Laboratory work sodium (mmol/l)[2] |
| 27 | `Creatinine` | Num | Laboratory work creatinine (umol/l)[2] |
| 28 | `Urea` | Num | Laboratory work blood urea (mmol/l)[2] |
| 29 | `CRP` | Num | Laboratory work CRP (mg/l)[2] |

Attributes that the consortium participants deemed relevant for the prediction of the 30 day rehospitalization problem.

[1]Num - numerical values; Cat - categorical values; CatL - List of categorical values.

[2]Laboratory tests from serum - value of the last test in that admission
