## Appendix 2 Data descriptors for "Crowdsourcing AI solutions in early rehospitalisation as a pilot for healthcare regulatory sandbox"

### S2 Appendix Dataset features descriptors

In our rehospitalization challenge dataset, we observed 641 unique ICD-10 codes for admission diagnoses, reflecting initial clinical presentations. Through in-hospital diagnostic processes, these were refined to 323 unique ICD-10 discharge codes, representing more specific final diagnoses. The dataset encompassed 486 unique diagnoses overall, illustrating the convergence from broader admission to more precise discharge classifications.

Within the admission diagnose there is 16 unique rare diagnoses with total of 56 rows and within discharge diagnosis there is 12 unique rare diagnoses with total of 32 rows, including diseases such as Eisenmenger syndrome, Brugada syndrome, and Fallot's tetralogy. Due to the transformation of ICD−10 codes, there was no need to exclude these rows. Croatian Association for Rare Diseases has a database of 436 rare diseases in Croatia from The Croatian Association for Rare Diseases https://www.zivotorg.org/en/associations/croatian-association-for-rare-diseases/. To further protect patient identities, codes associated with rare diseases were replaced with the 'X00' code for `Admission`, `Discharge`, and `Secondary Diagnoses`.

The WHO is responsible for the development and maintenance of the Anatomical Therapeutic Chemical (ATC)[1] Classification System, which is globally recognized as the standard for drug classification. Substances used for medical purposes are stored in the database in form of ATC codes. For the purpose of the challenge, the WHO ATC classification system version 2023 was used [1], which the Croatian Health Insurance Fund modified by adding a special category labelled `99_NOT_LISTED_WHO` that includes a heterogeneous set of codes not listed by WHO but used in the Croatian healthcare system.

Laboratory results variables extracted from the hospital information system noted within the Table 1 [Appendix A]: `Glucose`, `Potassium`, `Sodium`, `Creatinine`, `Urea` and `CRP` had to be omitted because the data was available only for the last 2 years within the extracted features dataset.

**Table 1 Metadata for the challenge phase 1 synthetic data phase and phase 2 anonymised data SPE phase.**

| | Column/Columns | Type[1] | Description |
|---|---|---|---|
| 1 | `AdmissionDx` | Cat | Diagnosis at patient admission, encoded in ICD−10 code. |
| 2 | `AdmissionType` | Cat | Hospital admission, binary EMERGENCY or ELECTIVE. |
| 3 | `PreviousAdmissionDays` | Num | Number of days elapsed since the patient's last hospital treatment. |
| 4 | `LOS` | Num | Length of hospital stay in days. |
| 5 | `Age_Group` | Num | Age at discharge, in years, categorised into 5-year groups. |
| 6 | `Gender` | Cat | Patient gender, binary M, F. |
| 7 | `Surgery_Count` | Num | Number of surgical procedures during hospital stay. |
| 8 | `LOS_ICU` | Num | Number of days spent in the intensive care unit (ICU). |
| 9 | `Discharge_Specialty` | Cat | Number representing the clinic at discharge within the hospital (e.g., cardiology, ICU). |
| 10 | `Dx_Discharge` | Cat | Diagnosis indicated at patient discharge, encoded in ICD−10 codes.<br>Rare diseases encoding category - XX. |
| 11 | `Discharge_Status` | Cat | Discharge status after hospital treatment: HOME, DIED (NOT EXAMINED),<br>OTHER, TREATMENT TERMINATED AGAINST DOCTOR'S ADVICE, ... |
| 12 | `Weigth_Discharge` | Num | Last recorded weight of the patient during the hospitalization. |
| 13 | `Height_Discharge` | Num | Last recorded height of the patient during the hospitalization. |
| 14 | `Education` | Cat | Socioeconomic level of education (e.g. HIGHER EDUCATION COMPLETED). |
| 15 | `Current_Work_Status` | Cat | Work status (e.g. RETIRED, FULL-TIME, UNEMPLOYED/PARTY, UNKNOWN). |
| 16[2] | `Dx_Secondary` | CatL | List of patient comorbidities, encoded in ICD-10 codes. |
| 17[2] | `Administered_Drugs (ATK 7)` | CatL | Therapeutic drugs administered during hospitalization, recorded as a list<br>and encoded using ATC-7 (WHO classification) codes. |
| 18[3] | `Features_Drugs` | Num | Featurization of `Administered_Drugs` if not found in the WHO list of medications,<br>it is recorded in the column 99_NOT_LISTED_WHO. Columns range from<br>'99_NOT_LISTED_WHO' to 'zolpidem', there are a total of 237 (synth) variables<br>(columns); 303 (SPE) variables. Drug combinations are preserved as columns. |
| 19[3] | `Features_Dx_Secondary` | Num | Secondary diagnoses are mapped to ICD-10 chapter letters. Columns indicate the<br>frequency of occurrence of each chapter (e.g., diagnoses A23 and A10 is mapped to column A = 2).<br>There are 21 variables in the synthetic dataset and 25 in the original dataset. |
| 20 | `Label` | Cat | 0 - the patient was not readmitted within 30 days for hospital treatment after discharge,<br>1 - the patient was admitted to hospital treatment within 30 days. |

[1]Num - numerical values; Cat - categorical values; CatL - List of categrorical values;

[2]Data was distributed in phase 2 only within the secure processing environment (SPE)

[3]Entries have different number of columns between phase 1 and phase 2, during synthetizing of the data some columns had to be dropped making synthetic data even safer.
